# Prospective assessment of DCE-MRI parameters associated with advanced mandibular osteoradionecrosis after IMRT for head and neck cancer

**DOI:** 10.1101/2019.12.23.19015651

**Authors:** Joint Head and Neck Radiotherapy-MRI Development Cooperative, Abdallah S. R. Mohamed, Renjie He, Yao Ding, Jihong Wang, Joly Fahim, Baher Elgohari, Hesham Elhalawani, Andrew D. Kim, Hoda Ahmed, Jose A. Garcia, Jason M. Johnson, R. Jason Stafford, James A. Bankson, Mark S. Chambers, Vlad C. Sandulache, Clifton D. Fuller, Stephen Y. Lai

## Abstract

**Purpose:** We aim to characterize the quantitative DCE-MRI parameters associated with advanced mandibular osteoradionecrosis (ORN) compared to the contralateral normal mandible.

**Experimental Design:** Patients with the diagnosis of advanced ORN after curative-intent radiation treatment of head and neck cancer were prospectively enrolled after institutional-review board approval and study-specific informed consent. Eligibility criteria included; age>18 years, pathological evidence of head and neck malignancy with history of curative-intent external beam radiotherapy; patients with clinically confirmed high-grade ORN requiring surgical intervention; and no contraindications to MRI. The DCE-MRI acquisition consisted of a variable flip angle T1 mapping sequence and a multi-phase 3D FSPGR sequence. Quantitative maps generated with the Tofts and extended Tofts pharmacokinetic model were used for analysis. Motion correction was applied. Manual segmentation of advanced ORN 3-D volume was done using anatomical sequences (T1, T2, and T1+contrast) to create ORN volumes of interest (ORN-VOIs).

Subsequently, normal mandibular VOIs were segmented on the contralateral healthy mandible of similar volume and anatomical location (i.e., mirror image) to create self-control VOIs. Finally, anatomical sequences were co-registered to DCE sequences, and contours were propagated to the respective quantitative parameter maps.

**Results:** Thirty patients were included. Median age at diagnosis was 58 years (range 19-78), and 83% were men. The site of tumor origin was in the oropharynx, oral cavity, salivary glands, and nasopharynx in 13, 9, 6, and 2 patients, respectively. The median time to ORN development after completion of IMRT was 38 months (range 6-184). There were statistically significant higher K^trans^ and V_e_ values in ORN-VOIs compared with controls (0.23 vs. 0.07 min^−1^, and 0.34 vs. 0.15, p <0.0001 for both) using matched pairs analysis. The average relative increase of K^trans^ in ORN-VOIs was 3.2 folds healthy mandibular control VOIs. Moreover, the corresponding rise of V_e_ in ORN-VOIs was 2.7 folds higher than the controls. Using combined K^trans^ and V_e_ parameters, 27 patients (90%) had at least a 200% increase of either of the studied parameters in the ORN-VOIs compared with their healthy mandible control VOIs.

**Conclusion:** Our results confirm there is a quantitatively significant higher degree of leakiness in the mandibular vasculature as measured using DCE-MRI parameters of areas affected with an advanced grade of ORN versus healthy mandible. We were able to measure significant increases in quantitative metrics compared to values from the non-ORN mandibular bone. Further efforts are ongoing to validate these findings to enable the use of these DCE-MRI parameter thresholds for the early detection of subclinical cases of ORN.

## Introduction

Osteoradionecrosis (ORN) of the mandible is a debilitating complication of external beam radiation therapy (EBRT) for head and neck cancer patients.^1-3^ Head and neck squamous cell carcinoma (HNSCC) has an estimated annual incidence of approximately 62,000 cases in the United States.^4^ The incidence of Human Papillomavirus (HPV) associated HNSCC continues to rise unabated and is expected to continue to rise for the next two decades until the effects of immunization will begin to impact incidence.^5,6^

At our institution, the rate of ORN development in HNSCC patients following EBRT is approximately 7% despite aggressive dental care and close follow up.^7^ Even with a relatively low incidence, the prevalence and burden of ORN is expected to rise because of the excellent prognosis for HPV+ patients (i.e. 5-year overall survival of 80-90% for most patients). Therefore, it is expected that the US will accumulate a large population of adults with a history of mandibular radiation, a nearly 10-fold increase compared to historical trends when prevalence was lower due to higher mortality of non HPV-associated cancers.

Despite the use of more conformal EBRT techniques such as intensity-modulated radiotherapy (IMRT), the mandible remains exposed to significant radiation doses because of its close proximity to target volumes that can eventually lead to the development of ORN especially when coupled with infection and/or dental manipulation.^7,8^ Early-stage ORN can be controlled with conservative measures such as antibiotics, surgical debridement, hyperbaric oxygen (HBO) therapy, pentoxifylline or tocopherol.^9,10^ However, progression to advanced ORN typically requires extensive surgical resection and complex reconstruction and leads to a substantial reduction in the quality of life of HNSCC survivors.^11,12^

Anatomic imaging using CT or conventional MRI does not identify ORN-related bony changes until relatively late in the process, when the patient is generally already experiencing symptoms. ^13,14^ Dynamic contrast-enhanced magnetic resonance imaging (DCE-MRI) is a clinically available imaging method that was shown to detect early-stage idiopathic osteonecrosis of the femur otherwise not visible on conventional MRI.^15^ DCE-MRI parameters can be used to monitor bone-healing secondary to trauma or fracture, as well as chronic changes in bone health associated with age-related osteoporotic changes.^16-18^

The most commonly accepted biological mechanism of ORN development remains that summarized by Marx’s three H’s of hypoxic, hypovascular, and hypocellular tissue. Therefore, we expect that shifts in vascularity may portend development of ORN.^19^ Hence, we focused on DCE-MRI as opposed to other imaging modalities. Our group has recently demonstrated that DCE-MRI can be used to detect alterations in bone vascularity following definitive radiotherapy to head and neck cancer patients.^20^ However, we do not yet know how early changes in bone vascularity during radiation correlate with subsequent development of ORN. In order to develop a predictive, imaging based biomarker of ORN development, it is therefore critical to identify DCE-MRI parameters in patients with existing ORN. This will facilitate the discrimination of the quantitative DCE-MRI parameters associated with injured versus healthy mandibular subregions. This characterization will ultimately serve as a guide to monitor temporal DCE-MRI changes following EBRT in attempt to early detect mandibular pathology before the development of symptoms.

Based on existing clinical and preclinical data, we hypothesized that ORN is associated with critical changes in bone vascularity reflected in common DCE MRI parameters namely V_e_ and Ktrans, as a reflection of overall poor vascular flow and integrity. To this end, we sought to characterize the quantitative DCE-MRI parameters associated with the established diagnosis of advanced mandibular ORN compared with normal mandible in the context of a prospective clinical study with high intrinsic imaging acquisition consistency.

## Methods

### Patient selection

Patients with a confirmed diagnosis of advanced ORN developed after curative-intent radiation treatment of head and neck cancer were prospectively enrolled in an observational imaging study (NCT03145077) after institutional-review board approval and study-specific informed consent. Eligibility criteria included age>18 years, pathological evidence of head and neck malignancy with history of curative-intent external beam radiotherapy, patients with clinically confirmed high-grade ORN requiring surgical intervention, good performance status (ECOG score 0-2), and no contraindications to MRI. Clinical staging of ORN was conducted using the Common Terminology Criteria for Adverse Events (CTCAE) version 4.0.

### DCE-MRI Imaging

DCE-MRI scans were obtained using GE 3.0T Discovery MR750 scanners with a 6-channel Flex phased-array coil (GE Healthcare Technology, Milwaukee, WI). Prior to DCE-MRI, T1 mapping was performed using six variable flip angles (FA) 3D spoiled gradient recalled echo (SPGR) sequence (FA□=□2°, 5°, 10°, 15°, 20°, and 25°; TR/TE□=□5.5/2.1□ms, FOV = 25.6 cm, slice number = 30, voxel size □=□2□×□2□×□4□mm^3^). The DCE-MRI was acquired using a multi-phase 3D Fast SPGR sequence to gain sufficient signal-to-noise ratio (SNR), contrast, and temporal resolution (FA□=□15°, TR/TE□=□3.6/1□ms, voxel size = 2×2×4□mm3, temporal resolution□=□5.5s, number of repetitions□=□56, pixel bandwidth□=□326□Hz, ASSET acceleration= 2). Gadobutrol (Gadovist, Bayer Healthcare, Germany) was administered at a dose of 0.1 mmol/kg of body weight at 3 ml/s followed by the same amount of saline at 3 ml/s, via a power injector (Spectris MR Injector, MedRad, Pittsburgh Pa).

### Computation of the kinetic model

Post-processing of the DCE-MRI images was performed at a workstation running in-house Matlab based pipeline (Matlab, MathWorks, MA, USA). Before quantitative analysis, motion correction and noise suppression were applied using simultaneous spatial and temporal higher-order total variations regularizations (HOTVs) as described by Chan et al.^21^ As shown in Figure 1, filtering with HOTVs demonstrated noise suppression as well as motion reduction. To quantify the physiological parameters using DCE-MRI, the arterial input function (AIF) of the contrast agent (CA) entering the tissue was pre-determined. T1 map was calculated to convert the signal intensity into concentration time course. Extended Tofts model assumes that the CA resides in and exchanges between two compartments in the tissue: the vascular space and extracellular extravascular space (EES). When this model is used, the differential equation describing the kinetic behavior of the CA in the tissue of interest is given by:

**Figure 1.**
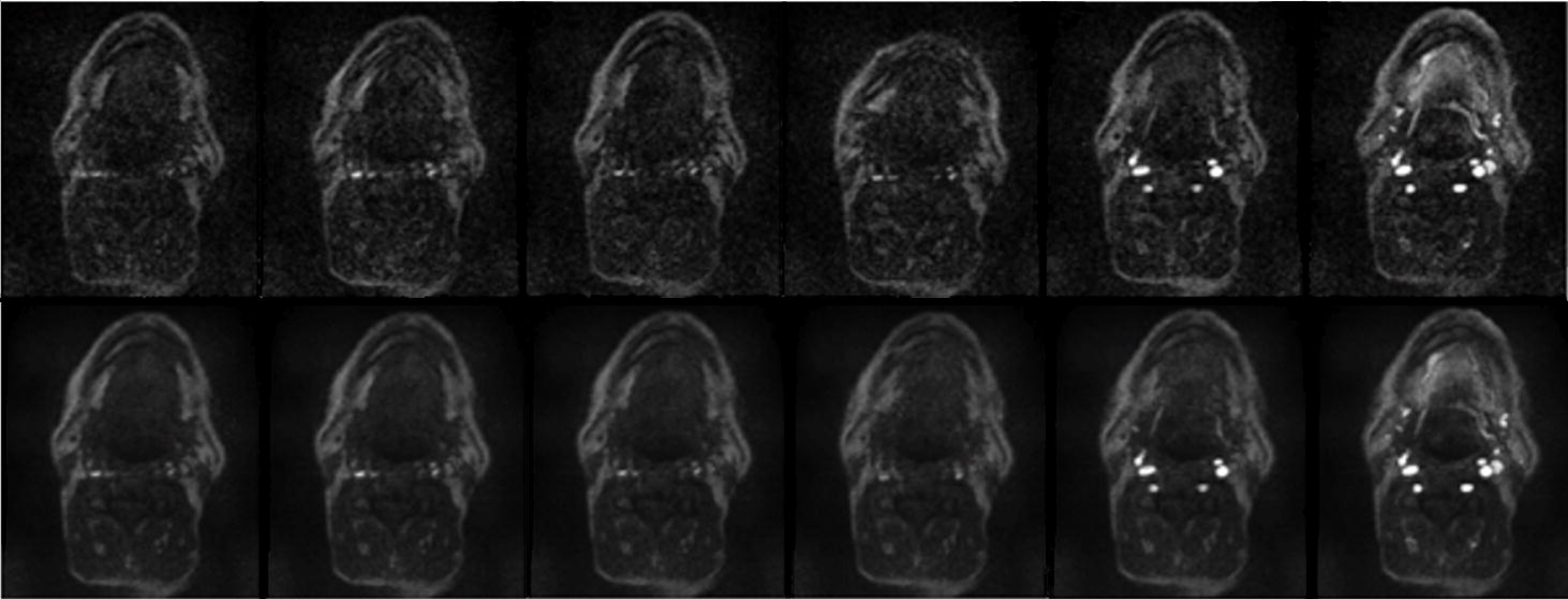
Consecutive DCE MRI frame time series (from left to right) before (row up) and after (row down) filtering with HOTVs.

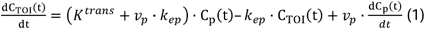

where *C*_*TOI*_(*t*) and *C*_*p*_(*t*) are the concentrations of the CA at time *t* in the tissue of interest (TOI) and blood plasma, respectively, *K*^*trans*^ are the transfer (CA permeability rate) constants between blood plasma and the EES of the TOI. *k*_*ep*_ = *K*^trans^/*V*_*e*_ is the transfer (CA permeability rate) constant (min^−1^) from the TOI back to the blood plasma, where *V*_*e*_ is the distribution volume fractions of CA in the EES per unit volume of tissue. When the kinetic model includes a vascular term, *V*_*p*_ that is the capillary plasma volume fractions per unit volume of tissue.Otherwise, by ignoring vascular term, the extended Tofts model is reduced to Tofts model as:

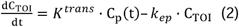

The AIF for the pharmacokinetic (PK) analysis was derived from selected arteries. The relevance of several analytical AIF models in DCE-MRI have been previously extensively investigated.^22^ Recently computer simulations were performed to evaluate and compare a population of AIF models with the Parker model.^23^ The results demonstrated that a six-parameter linear function plus bi-exponential function AIF model was almost equivalent to the Parker AIF. It should be noted the former is computationally faster and more reliable in functional fitting when compared to the Parker AIF. However, predetermining the arrival time (AT) and time to peak (TTP) of upslope for each AIF time series is usually not accurate in using the above six-parameter model. Therefore, we extended the six-parameter model to a bi-exponential and bi-linear function where the AT and TTP of the upslope are included as parameters to be estimated in AIF fitting. The new AIF model function (AIFM) was defined as:

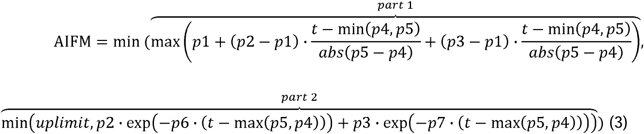

where *p*1 to *p*7 are parameters to be determined by functional fitting^23^, *min*(*p*4,*p*5) is AT, and *max*(*p*4,*p*5) is TTP; *t* is time points, *min* is minimal value operator, *max* is the maximal value operator, and *abs* is absolute value operator. “*uplimit”* is a constant that is estimated by the maximal possible value of data to be fitted. The fitting cost function is implemented by optimization on ‖DATA - AIFM‖, where DATA could be DCE concentration or DCE signal. By using this extended AIF model function, the AIF fitting can be completed in a more precise and reliable manner. Figure 2 shows the fitting process as well as the fitting results where a 56 time point AIF time series is presented. The PK modelling was done on a pixel-by-pixel basis using a linearization equation of the models used (i.e. Tofts and extended Tofts) as described by Kenya Murase.^25^ Subsequently, we implemented the linear least squares method to acquire the PK parameters.^25,26^ This method is preferred to the conventional nonlinear least squares method^24^, because it is faster and it does not require initial estimation, and has no local optima problems.

**Figure 2.**
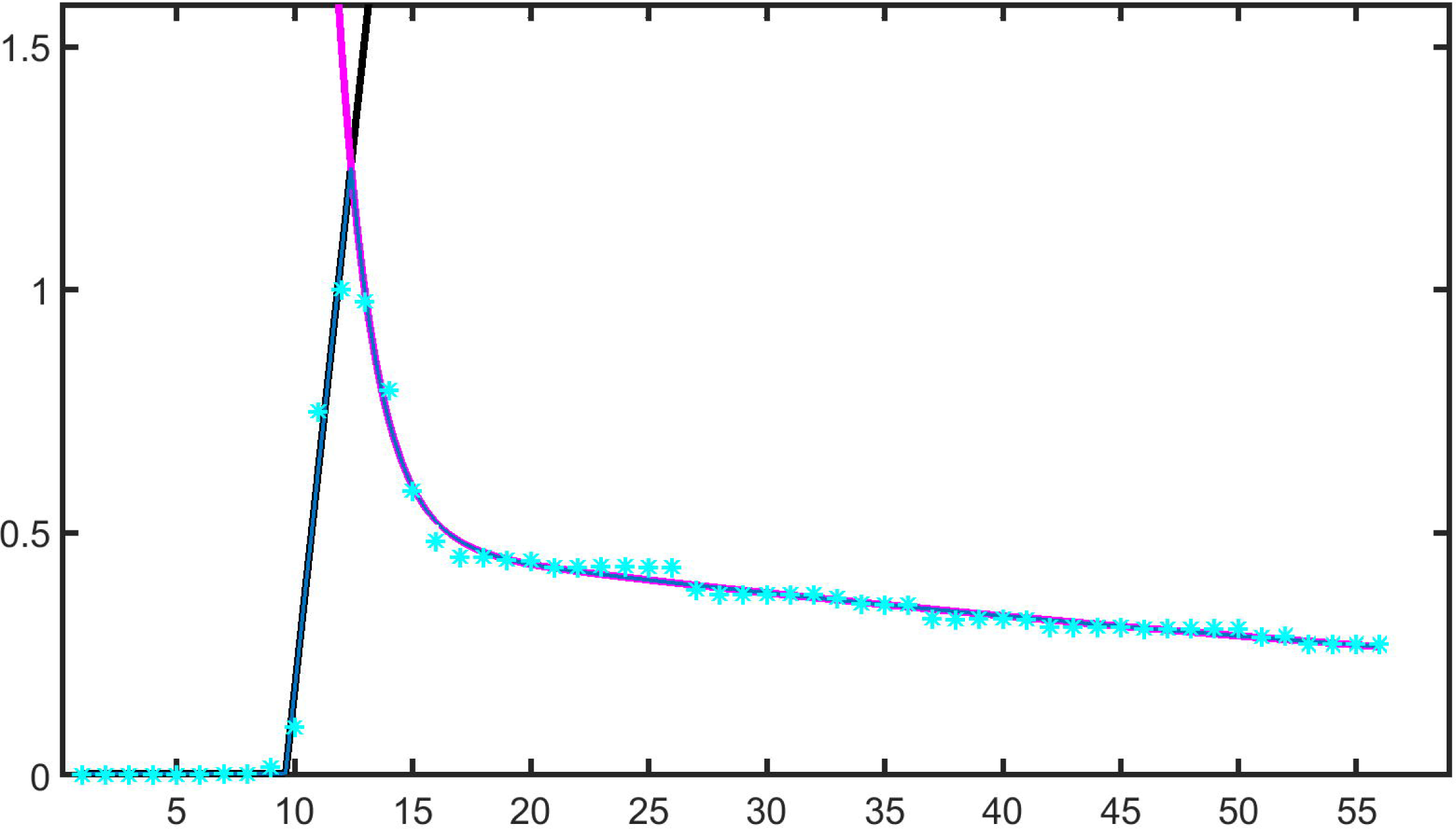
The fitting process and fitting results where a 56 time point AIF time series (in Cyan) is presented.

### Image segmentation and registration

Manual segmentation of mandibular volumes harboring ORN was done by an expert radiation oncologist (ASRM) and reviewed by expert neuroradiologist (JMJ). The segmentation was done using the MRI anatomical sequences (T1, T2, and T1+contrast) as well as co-registered contrast enhanced CTs to create ORN volumes of interest (ORN-VOIs) for all included patients. The segmentation included abnormal signal intensity or irregular gadolinium enhancement of bone marrow and soft tissues seen on MRIs^27,28^ as well as cortical erosions, sequestrations, and/or fractures seen on CTs^29^. Subsequently, normal mandibular VOIs were created on the contralateral healthy mandible of similar volume and anatomical location (i.e. mirror image) to create self-control VOIs. Finally, the MRI anatomical sequences were co-registered to the DCE-MRI sequences and then the contours were propagated to the respective quantitative parameter maps. This workflow is graphically summarized in Figure 3.

**Figure 3.**
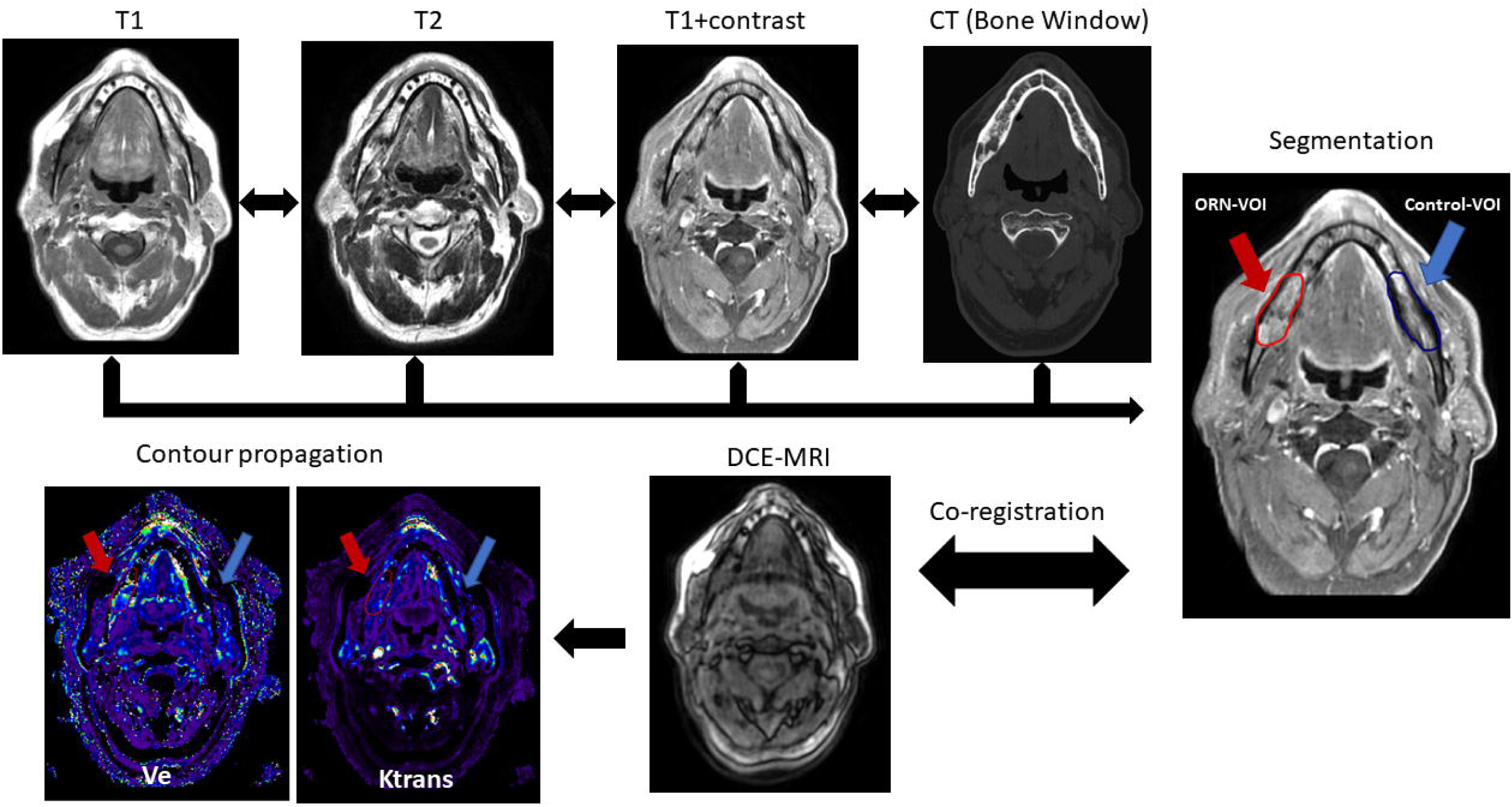
Workflow of advanced ORN analysis.

### Statistical analysis

Continuous data were presented as mean ± standard deviation, while categorical data were presented as proportions. The comparison of quantitative DCE-MRI parameters between ORN and Control VOIs was assessed using the non-parametric Wilcoxon signed-rank test. The effect size was calculated and the magnitude of the effect size was determined using Cohen’s criteria where r of 0.1=small effect, 0.3=medium effect and 0.5=large effect.^30^ P values <0.05 were deemed statistically significant. All statistical analyses were performed using JMP 14 Pro (SAS Institute, Cary, NC).

## Results

### Patients

Thirty patients with grade 3 ORN requiring surgery were included. Median age at diagnosis was 58 years (range 19-78), and 83% were men. The site of tumor origin was in the oropharynx, oral cavity, salivary glands, and nasopharynx in 13, 9, 6, and 2 patients, respectively. IMRT was the radiation technique for all patients. The median IMRT prescription dose was 68 Gy in 32 fractions. The median time to ORN development after completion of IMRT was 38 months (range 6-184). Table 1 summarizes patient, disease, and treatment criteria.

**Table 1.**
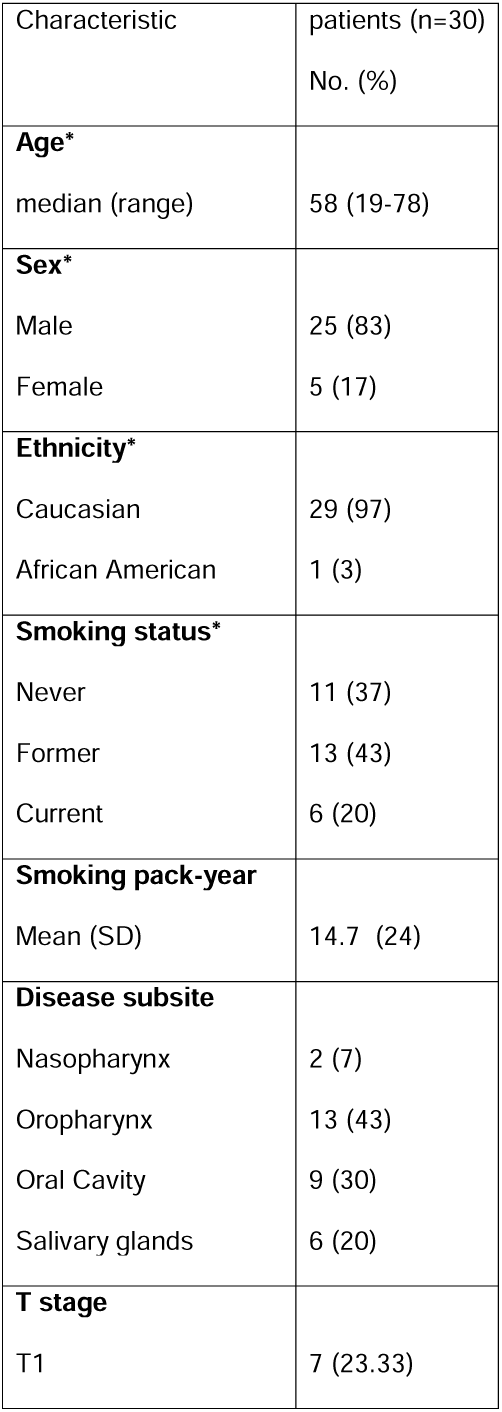
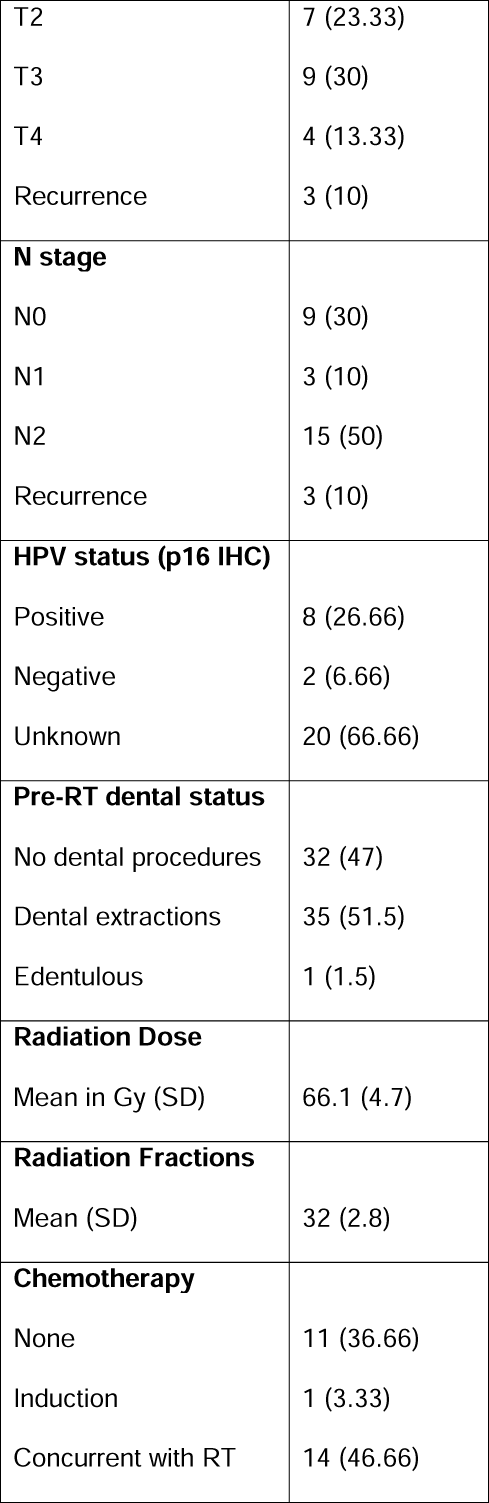
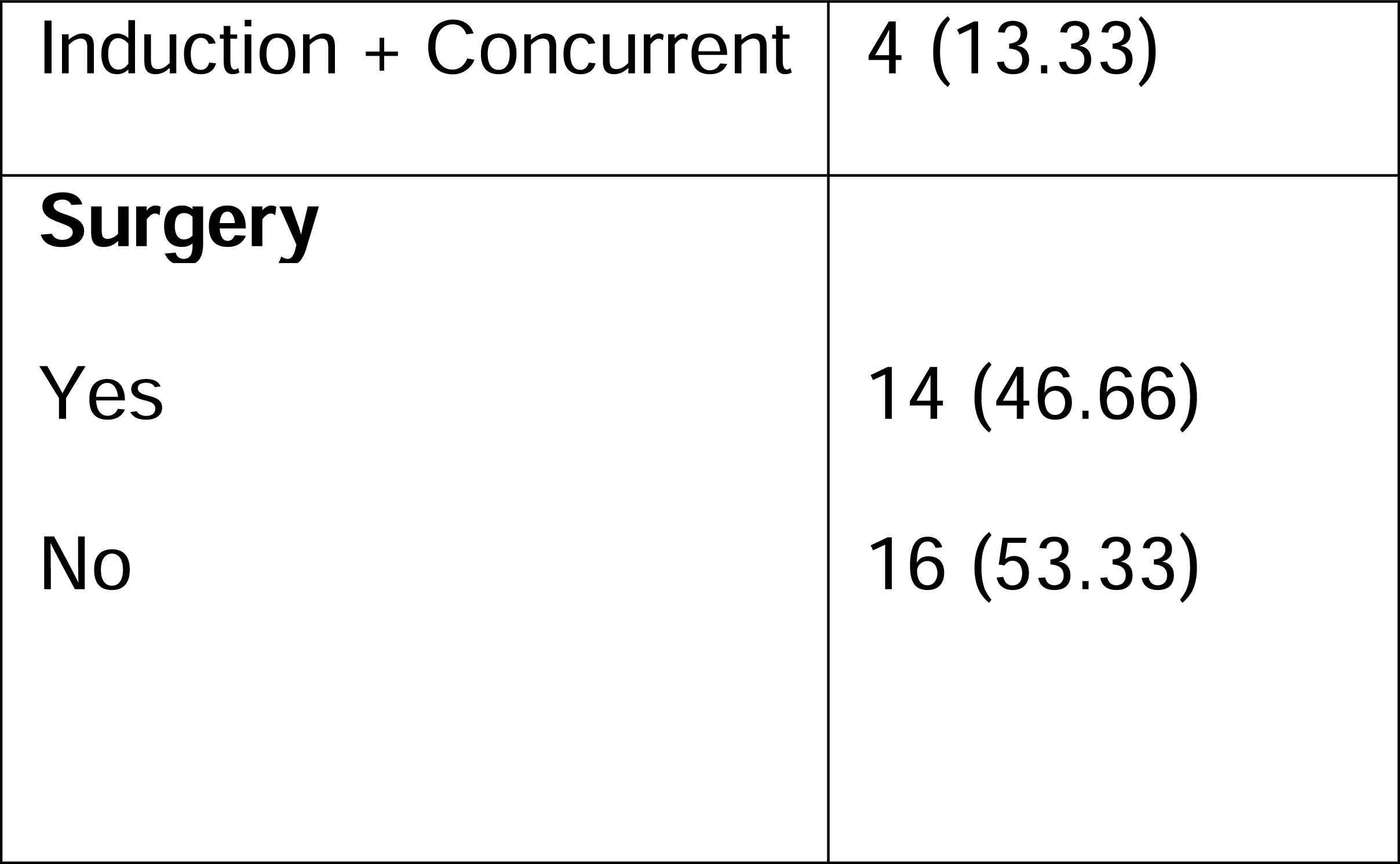
Patient, disease, and treatment characteristics

| Characteristic | patients (n=30)<br>No. (%) |
| --- | --- |
| **Age\*** |  |
| median (range) | 58 (19-78) |
| **Sex\*** |  |
| Male | 25 (83) |
| Female | 5 (17) |
| **Ethnicity\*** |  |
| Caucasian | 29 (97) |
| African American | 1 (3) |
| **Smoking status\*** |  |
| Never | 11 (37) |
| Former | 13 (43) |
| Current | 6 (20) |
| **Smoking pack-year** |  |
| Mean (SD) | 14.7 (24) |
| **Disease subsite** |  |
| Nasopharynx | 2 (7) |
| Oropharynx | 13 (43) |
| Oral Cavity | 9 (30) |
| Salivary glands | 6 (20) |
| **T stage** |  |
| T1 | 7 (23.33) |
| T2 | 7 (23.33) |
| T3 | 9 (30) |
| T4 | 4 (13.33) |
| Recurrence | 3 (10) |
| **N stage** |  |
| N0 | 9 (30) |
| N1 | 3 (10) |
| N2 | 15 (50) |
| Recurrence | 3 (10) |
| **HPV status (p16 IHC)** |  |
| Positive | 8 (26.66) |
| Negative | 2 (6.66) |
| Unknown | 20 (66.66) |
| **Pre-RT dental status** |  |
| No dental procedures | 32 (47) |
| Dental extractions | 35 (51.5) |
| Edentulous | 1 (1.5) |
| **Radiation Dose** |  |
| Mean in Gy (SD) | 66.1 (4.7) |
| **Radiation Fractions** |  |
| Mean (SD) | 32 (2.8) |
| **Chemotherapy** |  |
| None | 11 (36.66) |
| Induction | 1 (3.33) |
| Concurrent with RT | 14 (46.66) |
| Induction + Concurrent | 4 (13.33) |
| **Surgery** |  |
| Yes | 14 (46.66) |
| No | 16 (53.33) |

### DCE-MRI parameters

The median volume of segmented VOIs was 5.2 cm^3^ (range 1.8-10.9). Using the extended Tofts model, the average K^trans^ values in ORN-VOIs were significantly higher compared with controls (0.23±0.25 vs 0.07±0.07 min^−1^, p<0.0001). The average relative increase of K^trans^ in ORN-VOIs was 3.2 folds those the healthy mandibular control VOIs (range 1.2-10.3). The effect size was large with r=0.52.

Likewise, the average V_e_ values in ORN-VOIs was significantly higher compared with controls (0.34±0.27 vs 0.15±0.15, p<0.0001). The average relative increase of V_e_ in ORN-VOIs was 2.7 folds those the healthy mandibular control VOIs (range 1.2-6.9). The effect size was also large with r=0.69.

Using combined K^trans^ and V_e_ parameters, 27 patients (90%) displayed at least double the increase of either of the studied parameters in the ORN-VOIs compared with their healthy mandible control VOIs. Figure 4 depicts the comparison of K^trans^ and V_e_ values in ORN-VOIs compared with controls.

**Figure 4.**
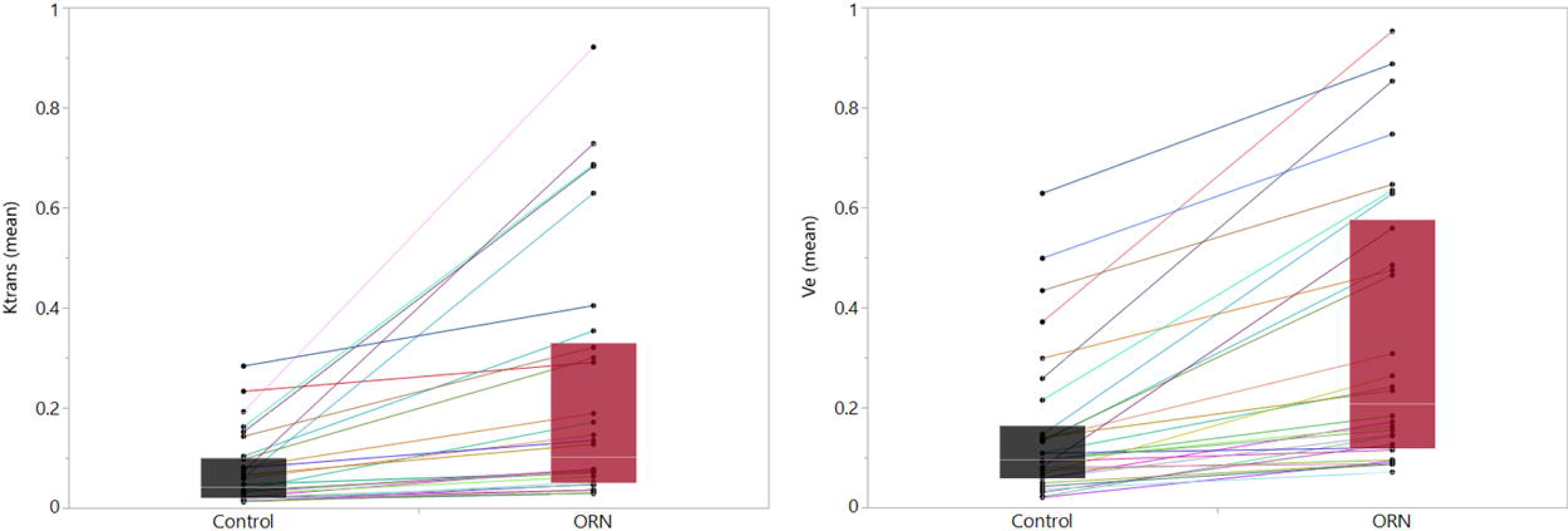
Boxplots shows a statistically significant higher K^trans^ and V_e_ values in ORN-VOIs compared with controls.

V_p_ was also significantly higher in ORN-VOIs compared with controls (0.17±0.2 vs 0.07±0.12, p=0.008). However, K_ep_, as expected, did not show a significant difference between ORN-VOIs versus controls (0.46±0.19 vs 0.5±0.13 min^−1^, p=0.1) because of the increase of both K^trans^ and V_e_ parameters.

Using the Tofts model, the mean K^trans^ values in ORN-VOIs were, likewise, significantly higher compared with controls (0.27±0.29 vs 0.08±0.08 min^−1^, p<0.0001). The average relative increase of K^trans^ in ORN-VOIs was 3.4 folds of the control VOIs (range 1.5-12.3). In addition, the mean V_e_ values in ORN-VOIs was significantly higher compared with controls (0.34±0.28 vs 0.13±0.13, p<0.0001). The average relative increase of V_e_ in ORN-VOIs was 4.04 folds of the healthy mandibular control VOIs (range 1.2-15.3). Using combined K^trans^ and V_e_ parameters, also showed that 90% of patients had more than two folds increase of either of the studied parameters in the ORN-VOIs compared with their healthy mandible control VOIs.

## Discussion

Nearly 4 decades ago, R.E. Marx postulated a theory for development of ORN predicated in large part on altered bone vascularity, resulting in poor regenerative capacity and a decreased ability to resist mechanical and microbial insults.^19^ Although this remains the most likely mechanism for ORN, to date scientists and clinicians have lacked the means to study bone vascularity with sub-centimeter spatial resolution in a non-invasive manner and have largely been forced to infer mechanisms of ORN development by combining limited pre-clinical studies, with static anatomic imaging and histologic evaluation of ORN specimens. For the first time, we now have the opportunity to leverage a clinically available imaging approach to provide real-time, non-invasive information about bone vascularity in the context of ORN (Figure 5). This represents a breakthrough both in our ability to study this devastating disease and to begin to develop clinical trials design to ameliorate the disease using objective, quantitative measures.

**Figure 5.**
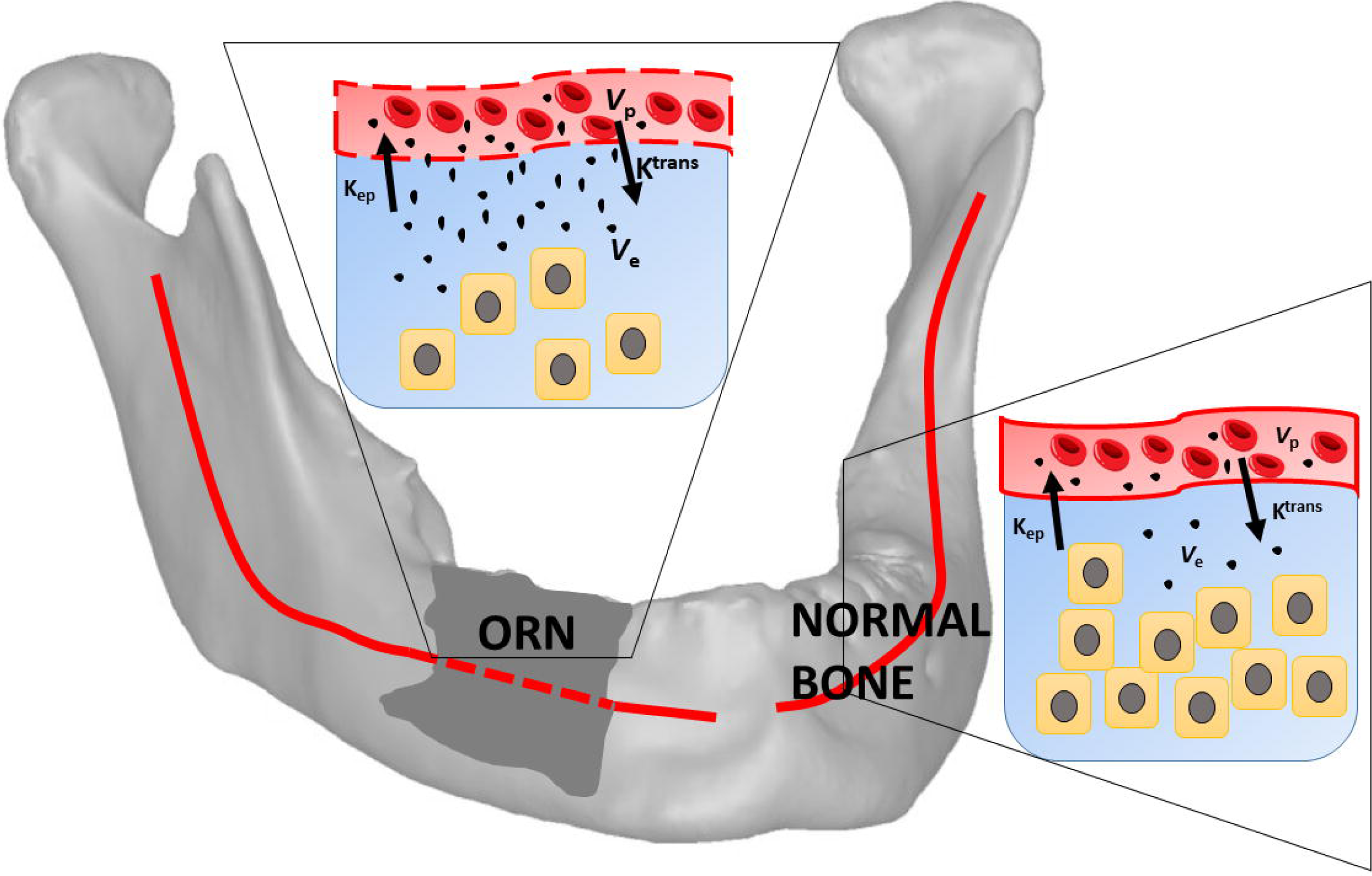
Schematic cartoon of the suggested mechanistic changes of ORN.Right mandibular body with area of ORN and associated altered vascularity; contralateral body with normal body and vascularity. Normal blood vessels have intact walls (continuous lines) with well-regulated fractional volume plasma (V_p_) and appropriate exchange across the vessel wall (Ktrans) to extracellular extravascular compartment (V_e_) and normal cellularity. Vessels associated with ORN demonstrate higher V_p_ with increased leakiness (Ktrans) through fragile vessels (dotted lines) that leads to increased V_e_ and hypocellularity.

Our findings demonstrated a distinct profile of DCE-MRI parameter maps in mandibular volumes harboring ORN as compared to the normal mandible. DCE parameters indicating vascular compromise showed a significantly higher degree of leakiness in the mandibular vasculature as measured using K^trans^ and V_e_ of areas affected with advanced grade of ORN versus healthy mandible. The fractional volume of plasma (V_p_) was also higher in ORN-ROIs. We were able to measure significant increases in quantitative parameters with an average increase of approximately three-fold of both K^trans^ and V_e_ compared to values from the healthy mandibular bone. The vast majority of patients (90%) had at least doubling of the values of either K^trans^ or V_e_ for ORN-VOIs as compared with control VOIs.

The quantitative perfusion characteristics of mandibular injury after radiation treatment of the head and neck cancer have never, to our knowledge, been assessed before. One study has previously investigated the qualitative nature of contrast enhancement of DCE-MRI in patients with mandibular ORN.^27^ That study showed that all patients with ORN had marked contrast enhancement of the osteoradionecrotic bone marrow, which was reduced after treatment with HBO treatment.^27^ DCE MRI has also shown the ability to detect early-stage idiopathic osteonecrosis of the femur not otherwise visible on conventional MRI as reported by Chan et al.^15^ In addition, DCE-MRI parameters were also used to monitor bone healing secondary to trauma or fracture, as well as chronic changes in bone health associated with age-related osteoporotic changes.^16-18^ DCE-MRI was also reported to identify changes related to the development of bony metastasis and tumor response to treatment.^31,32^

We have recently demonstrated that DCE-MRI can be used to detect radiation-induced changes in mandibular bone vascularity and showed dose-dependent changes in both K^trans^ and V_e_ in a subset of patients.^20^ Unlike the finding of our previous study that showed variability in the dose-dependent changes of vascular parameters where a percentage of patients had a decrease in the measured parameters after treatment, the current study only demonstrated an increase of these parameters. The increased fractional plasma and vascular leakiness in ORN areas reflects fragile vasculature that may be attributed to a process of neovascularization following the post-radiation chronic hypoxia in high-dose areas. A serial imaging study of the natural history of vascular changes of the mandible is currently ongoing to determine at which time point this phenomenon of neovascularization begins to develop and to what extent this development could be correlated with early ORN symptoms development. However, our findings suggest that a two-fold increase in either K^trans^ or V_e_ parameters is an alarming sign of ORN development if detected in patients with otherwise clinically apparent normal mandible after radiation treatment especially in areas exposed to higher doses of radiation due to tumor proximity.

Our group and others have previously demonstrated the dependency of DCE-MRI quantitative output on the nature of PK models used for analysis.^33,34^ Thereby, we used the most widely accepted models such as Tofts and extended Tofts models. We also used the patients’ contralateral mandible as an internal control to alleviate this model dependency. Additionally, we have recently shown, using multi-institutional comparison of patient-derived DCE-MRI data that quantitative values may not be reliably compared across different patients due to the difference in patient’s specific imaging parameters, pre-processing, and post-processing factors.^33^ Because of such limitation, in the current study we did not use the absolute values of the parameters associated with ORN but instead compared the relative changes of these parameters to respective controls in the same image for each patient using the contralateral healthy mandibular VOIs. Therefore, our results may be more reproducible and generalizable as they represent the relative changes measured in the irradiated mandibular areas compared with the normal non-irradiated bony area and hence we avoided the inter-subject variability of the parameter absolute values.

Despite that, the thirty patients included in this study may be perceived as a limited sample size. However, to date this represents the largest prospective quantitative imaging study of ORN ever reported. Furthermore, this study represents the initial characterization of quantitative vascular parameters driven from DCE-MRI for head and neck cancer patients treated with IMRT and affected by radiation-induced advanced ORN toxicity.

## Conclusion

Our results confirmed higher degree of vascular leakiness in the mandibular areas of ORN as measured using DCE-MRI parameters as compared with healthy mandible. Additional efforts will be required to develop DCE-MRI parameter into viable non-invasive biomarkers useful for the early detection of subclinical cases of ORN.

## Data Availability

De-identified data is available upon request.

## References

1. Teng MS, Futran ND. Osteoradionecrosis of the mandible. Current Opinion in Otolaryngology & Head and Neck Surgery. 2005;13(4):217–221.

2. Jereczek-Fossa BA, Orecchia R. Radiotherapy-Induced Mandibular Bone Complications. Cancer Treatment Reviews. 2002;28(1):65–74.

3. Sciubba JJ, Goldenberg D. Oral complications of radiotherapy. The Lancet Oncology. 2006;7(2):175–183.

4. Siegel RL, Miller KD, Jemal A. Cancer statistics, 2019. CA: A Cancer Journal for Clinicians. 2019;69(1):7–34.

5. O’Sullivan B, Huang SH, Su J, et al. Development and validation of a staging system for HPV-related oropharyngeal cancer by the International Collaboration on Oropharyngeal cancer Network for Staging (ICON-S): a multicentre cohort study. Lancet Oncol. 2016;17(4):440–451.

6. Chaturvedi AK, Anderson WF, Lortet-Tieulent J, et al. Worldwide trends in incidence rates for oral cavity and oropharyngeal cancers. J Clin Oncol. 2013;31(36):4550–4559.

7. Mohamed ASR, Hobbs BP, Hutcheson KA, et al. Dose-volume correlates of mandibular osteoradionecrosis in Oropharynx cancer patients receiving intensity-modulated radiotherapy: Results from a case-matched comparison. Radiotherapy and Oncology. 2017;124(2):232–239.

8. Beadle BM, Liao KP, Chambers MS, et al. Evaluating the impact of patient, tumor, and treatment characteristics on the development of jaw complications in patients treated for oral cancers: a SEER-Medicare analysis. Head & neck. 2013;35(11):1599–1605.

9. Costa DA, Costa TP, Netto EC, et al. New perspectives on the conservative management of osteoradionecrosis of the mandible: A literature review. Head Neck. 2016;38(11):1708–1716.

10. Lambade PN, Lambade D, Goel M. Osteoradionecrosis of the mandible: a review. Oral Maxillofac Surg. 2013;17(4):243–249.

11. Zaghi S, Miller M, Blackwell K, Palla B, Lai C, Nabili V. Analysis of surgical margins in cases of mandibular osteoradionecrosis that progress despite extensive mandible resection and free tissue transfer. Am J Otolaryngol. 2012;33(5):576–580.

12. Wang CC, Cheng MH, Hao SP, Wu CC, Huang SS. Osteoradionecrosis with combined mandibulotomy and marginal mandibulectomy. Laryngoscope. 2005;115(11):1963–1967.

13. Hamilton JD, Lai SY, Ginsberg LE. Superimposed infection in mandibular osteoradionecrosis: diagnosis and outcomes. J Comput Assist Tomogr. 2012;36(6):725–731.

14. Khojastepour L, Bronoosh P, Zeinalzade M. Mandibular bone changes induced by head and neck radiotherapy. Indian J Dent Res. 2012;23(6):774–777.

15. Chan WP, Liu YJ, Huang GS, et al. Relationship of idiopathic osteonecrosis of the femoral head to perfusion changes in the proximal femur by dynamic contrast-enhanced MRI. AJR Am J Roentgenol. 2011;196(3):637–643.

16. Dyke JP, Lazaro LE, Hettrich CM, Hentel KD, Helfet DL, Lorich DG. Regional analysis of femoral head perfusion following displaced fractures of the femoral neck. J Magn Reson Imaging. 2015;41(2):550–554.

17. Ma HT, Griffith JF, Zhao X, Lv H, Yeung DK, Leung PC. Relationship between marrow perfusion and bone mineral density: a pharmacokinetic study of DCE-MRI. Conference proceedings : Annual International Conference of the IEEE Engineering in Medicine and Biology Society IEEE Engineering in Medicine and Biology Society Annual Conference. 2012;2012:377–379.

18. Ma HT, Lv H, Griffith JF, Yuan J, Leung PC. Bone marrow perfusion of proximal femur varied with BMD--a longitudinal study by DCE-MRI. Conference proceedings : Annual International Conference of the IEEE Engineering in Medicine and Biology Society IEEE Engineering in Medicine and Biology Society Annual Conference. 2013;2013:2607–2610.

19. Marx RE. Osteoradionecrosis: a new concept of its pathophysiology. J Oral Maxillofac Surg. 1983;41(5):283–288.

20. Joint H, Neck Radiotherapy MRIDC, Sandulache VC, et al. Dynamic contrast-enhanced MRI detects acute radiotherapy-induced alterations in mandibular microvasculature: prospective assessment of imaging biomarkers of normal tissue injury. Scientific Reports. 2016;6:29864.

21. Chan T, Marquina A, Mulet P. High-order total variation-based image restoration. Siam J Sci Comput. 2000;22(2):503–516.

22. Balvay D, Ponvianne Y, Claudon M, Cuenod CA. Arterial input function: Relevance of eleven analytical models in DCE-MRI studies. Paper presented at: 2008 5th IEEE International Symposium on Biomedical Imaging: From Nano to Macro; 14–17 May 2008, 2008.

23. He D, Xu L, Qian W, Clarke J, Fan X. A simulation study comparing nine mathematical models of arterial input function for dynamic contrast enhanced MRI to the Parker model. Australas Phys Eng Sci Med. 2018;41(2):507–518.

24. Sourbron SP, Buckley DL. Classic models for dynamic contrast-enhanced MRI. NMR in Biomedicine. 2013;26(8):1004–1027.

25. Murase K. Efficient method for calculating kinetic parameters using T1-weighted dynamic contrast-enhanced magnetic resonance imaging. Magnetic resonance in medicine. 2004;51(4):858–862.

26. Jones KM, Pagel MD, Cardenas-Rodriguez J. Linearization improves the repeatability of quantitative dynamic contrast-enhanced MRI. Magnetic resonance imaging. 2018;47:16–24.

27. Store G, Smith HJ, Larheim TA. Dynamic MR imaging of mandibular osteoradionecrosis. Acta Radiol. 2000;41(1):31–37.

28. García-Ferrer L, Bagán JV, Martínez-Sanjuan V, et al. MRI of mandibular osteonecrosis secondary to bisphosphonates. AJR American journal of roentgenology. 2008;190(4):949–955.

29. Hermans R, Fossion E, Ioannides C, Van den Bogaert W, Ghekiere J, Baert AL. CT findings in osteoradionecrosis of the mandible. Skeletal Radiol. 1996;25(1):31–36.

30. Cohen J. Statistical power analysis for the behavioral sciences. 2nd ed. Hillsdale, N.J.: L. Erlbaum Associates; 1988.

31. Bauerle T, Bartling S, Berger M, et al. Imaging anti-angiogenic treatment response with DCE-VCT, DCE-MRI and DWI in an animal model of breast cancer bone metastasis. Eur J Radiol. 2010;73(2):280–287.

32. Dafni H, Kim SJ, Bankson JA, Sankaranarayanapillai M, Ronen SM. Macromolecular dynamic contrast-enhanced (DCE)-MRI detects reduced vascular permeability in a prostate cancer bone metastasis model following anti-platelet-derived growth factor receptor (PDGFR) therapy, indicating a drop in vascular endothelial growth factor receptor (VEGFR) activation. Magnetic resonance in medicine. 2008;60(4):822–833.

33. Joint H, Neck Radiotherapy MRIDC. A Multi-Institutional Comparison of Dynamic Contrast-Enhanced Magnetic Resonance Imaging Parameter Calculations. Sci Rep. 2017;7(1):11185.

34. Galbraith SM, Lodge MA, Taylor NJ, et al. Reproducibility of dynamic contrast-enhanced MRI in human muscle and tumours: comparison of quantitative and semi-quantitative analysis. NMR Biomed. 2002;15(2):132–142.

